# Biallelic variants in *RNU2-2* cause the most prevalent known recessive neurodevelopmental disorder

**DOI:** 10.1101/2025.08.26.25334179

**Authors:** Daniel Greene, Rodrigo Mendez, Jon Lees, Mafalda Barbosa, Alessandro Bruselles, Luigi Chiriatti, Cecilia Mancini, Undiagnosed Diseases Network, Enrico Silvio Bertini, Devon E. Bonner, Thomas A. Cassini, Kimberly M. Ezell, Natalia Gomez-Ospina, Lynette Rives, Vandana Shashi, Rebecca C. Spillmann, Mohamed Wafik, Marco Tartaglia, Jonathan A. Bernstein, Andrew D. Mumford, Matthew T. Wheeler, Ernest Turro

## Abstract

We recently showed that mutations in *RNU4-2* and *RNU2-2*, two genes that are transcribed into small nuclear RNA (snRNA) components of the major spliceosome, are prevalent causes of dominant neurodevelopmental disorders (NDDs). By genetic association comparing 12,776 NDD cases with 56,064 controls, we now demonstrate the existence of a recessive form of *RNU2-2* syndrome that, in England, is even more common than the dominant form. We inferred log Bayes factors for dominant and recessive models of association of 14.0 and 18.2, respectively, and observed 17 rare variants with a posterior probability of pathogenicity conditional on recessive association >0.8. This conservative threshold identified 18 probands (all with unaffected parents) and five affected siblings, each carrying two alleles in trans at these variants. A relaxed threshold of >0.6 identified a further 13 candidate probands. We estimate that recessive *RNU2-2* syndrome accounts for 7–10% of families with a diagnosed recessive NDD, and is 36–62% as prevalent as the dominant *RNU4-2*-related disorder ReNU syndrome. We identified a further seven cases in five pedigrees in two replication collections. Cases are characterized by intellectual disability, global developmental delay and seizures. The variants are predicted to destabilize stem loops and binding domains of the U2-2 snRNA that contribute to spliceosome quaternary structure, intron recognition and catalytic function. Despite this, whole-blood derived RNA-seq data from three patients did not reveal splicing defects, in line with previous analogous observations for dominant *RNU2-2* syndrome.

---

The major spliceosome is composed of approximately 100 proteins and five classes of small nuclear RNA (snRNA) — U1, U2, U4, U5 and U6 — each of which is encoded by a set of paralogous genes^1,2^. We recently reported that variants in the paralogs *RNU4-2* and *RNU2-2* cause two of the most prevalent NDDs^3,4^. These findings were reported independently by other groups^5,6^, two of which also identified a third rarer NDD caused by variants in *RNU5B-1*^6,7^. Variants in snRNA genes of the minor spliceosome are also causal of rare syndromic disorders^8–13^. However, whereas the reported minor spliceosome disorders are recessive, all three published NDDs caused by variants in major spliceosomal snRNAs are dominant. Here, we show that variants in *RNU2-2* cause one of the most prevalent recessive NDDs in England, where the disorder is between 24% and 42% as prevalent as the dominant *RNU4-2*-related disorder ReNU syndrome, which is caused almost exclusively by *de novo* mutations. The prevalence in other countries is unknown but will likely be higher in cultures in which consanguineous partnerships are common. Even among non-consanguineous families, recessive *RNU2-2* syndrome has a greater prevalence than the dominant form. Crucially, due to the hereditary nature of this novel disorder, it is amenable to interventions such as preconception counseling or antenatal genetic diagnosis.

We identified the recessive form of *RNU2-2* syndrome through a joint statistical analysis of the 100,000 Genomes Project (100KGP) and the Genomic Medicine Service (GMS) data in the National Genomic Research Library (NGRL)^14^ from pedigrees in England with rare disorders. Following our previously described approach^3,15^, we applied the BeviMed genetic association method^16^ to compare rare variant genotypes in the 41,132 canonical transcript entries in Ensembl v104 having a biotype other than ’protein_coding’ between 12,776 unrelated, unexplained NDD cases and 56,064 unrelated participants without an NDD. As in our previous analysis^4^, only two genes with a posterior probability of association (PPA) > 0.5 emerged: *RNU4-2* and *RNU2-2* (**Fig. 1a**). Given *RNU2-2* is only 191bp long, we were able to use read-backed phasing^17^ to delineate individuals with two rare variants in cis and those with two rare variants in compound heterozygosity (i.e., in trans). After phasing, we re-ran BeviMed using an adaptation that accounts for the phasing information to obtain refined log Bayes factors for dominant and recessive modes of inheritance. This analysis demonstrated only weak evidence of a recessive disorder associated with *RNU4-2* (log Bayes factor = 1.3) but very strong evidence of a recessive disorder associated with *RNU2-2* (log Bayes factor = 18.2) (**Fig. 1b**).

**Fig. 1.**
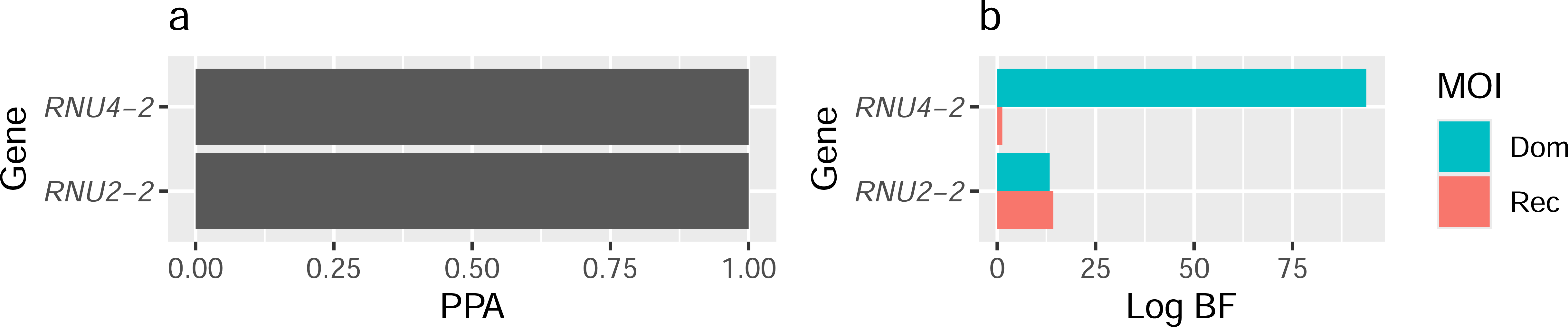
Genetic association results. **a**, BeviMed PPAs between each of *RNU4-2* and *RNU2-2* and case status. All other noncoding genes and pseudogenes had PPA < 0.5. **b**, For each of the two genes with PPA > 0.5, bars indicating BeviMed log Bayes factors with respect to the model representing a dominant (Dom) and the model representing a recessive (Rec) mode of inheritance (MOI).

For each rare variant in *RNU2-2*, we computed conditional probabilities of pathogenicity given the recessive inheritance model (PPP|rec). These probabilities indicate the degree of evidence supporting pathogenicity for each variant and depend on the patterns of homozygosity and compound heterozygosity in cases and controls. There were 18 unrelated cases (12 from the 100KGP and six from the GMS) with two alleles in trans at variants with PPP|rec > 0.8 (’Tier 1 variants’). We denoted these cases as high-confidence ’Tier 1 cases’ (**Fig. 2a**). Out of the 18 Tier 1 cases, 11 were homozygous and seven were compound heterozygous. In addition, there were 13 compound heterozygous cases who were not Tier 1 cases but were biallelic for variants with PPP|rec > 0.6. We labelled these lower confidence cases as ’Tier 2 cases’ and variants with 0.6 < PPP|rec < 0.8 as ’Tier 2 variants’ (**Extended Data** Fig. 1). None of the Tier 1 and Tier 2 variants were observed in homozygosity in gnomAD v4.

**Fig. 2.**
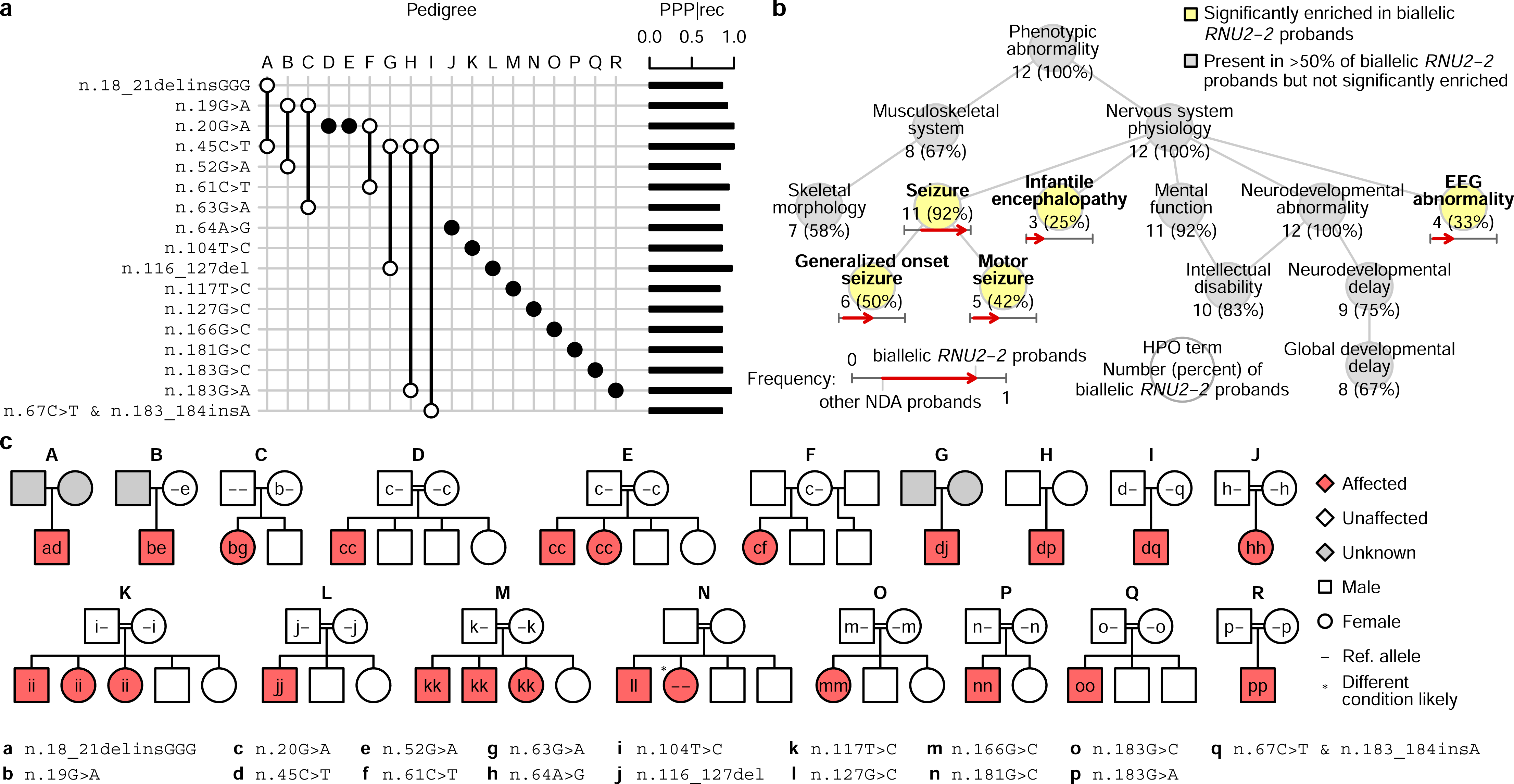
Tier 1 cases in the NGRL. **a**, Variant by pedigree grid illustrating the variants and genotypes corresponding to the Tier 1 cases. Filled circles denote homozygotes while empty circles denote heterozygotes. A pair of empty circles connected by a line denotes compound heterozygotes. The PPP|rec values for each variant are shown as bars on the right. **b**, Graph showing the ‘is-a’ relationships among HPO terms (abbreviated as needed for conciseness) present in at least half of the 12 Tier 1 cases in the discovery collection or significantly enriched among them relative to 9,112 probands with the ’Neurodevelopmental abnormality’ (NDA) HPO term in the 100KGP. The significantly overrepresented terms are highlighted. For each term, the number of cases with the term and the proportion that number represents out of 12 is shown. For each overrepresented term, the proportion of NDA-coded probands with the term and the proportion of Tier 1 *RNU2-2* cases in the 100KGP with the term are represented as the horizontal coordinate of the base and the head of an arrow, respectively. **c**, Pedigrees for biallelic Tier 1 cases in the discovery collection. Filled symbols indicate affected individuals. Double lines indicate consanguinity.

To corroborate the statistical association, we assessed the distinctiveness of the human phenotype ontology (HPO) terms assigned to the 12 Tier 1 cases in the 100KGP, omitting the GMS cases due to sparse HPO coding (**Extended Data** Fig. 2). The 12 cases were significantly more similar than expected by chance (*P*=0.001) (**Extended Data** Fig. 3). Statistically enriched terms relative to other NDD cases in the 100KGP included ’Generalized onset seizure,’ ’Motor seizure,’ ’Infantile encephalopathy’ and ’EEG abnormality’ (**Fig. 2b**). The terms ’Intellectual disability,’ ’Global developmental delay,’ and ’Abnormal skeletal morphology’ (primarily due to ’Microcephaly’) were attached to the majority of Tier 1 cases in the 100KGP but were not significantly enriched (**Fig. 2b**).

The 18 unrelated Tier 1 cases comprised seven compound heterozygotes and 11 homozygotes, the latter arising from consanguinity (**Fig. 2c**). In all cases where parental genotypes were available, the variants were inherited, not *de novo*. Cosegregation in five biallelic affected siblings across three families provided additional evidence for causality of n.20G>A, n.104T>C and n.116_127del. One affected sibling of a homozygous case with n.127G>C was homozygous wild type, but this did not contradict the likely pathogenicity of the variant because that sibling exhibited a different phenotype in keeping with trichothiodystrophy likely due to a separate homozygous variant in *GTF2H5*. Among the Tier 2 pedigrees, two affected siblings in two families inherited both variants in the corresponding probands (n.28C>G, n.158G>C, n.171_179del and g.62841593_62841624del) and there were no examples of non-cosegregation (**Extended Data** Fig. 1). All parents in both tiers were marked unaffected and had no assigned HPO terms.

To corroborate that variants in *RNU2-2* cause a recessive NDD, we examined the genomes of 5,323 participants from 1,759 families in the Undiagnosed Diseases Network (UDN) in the United States and of 220 unexplained NDD cases enrolled in the Ospedale Pediatrico Bambino Gesù Undiagnosed Patients Program (UPP) in Italy. In these collections, we identified five compound heterozygous probands and two affected siblings who were biallelic for rare variants in *RNU2-2* (**Fig. 3a**). Each case had at least one variant with strong support from the NGRL analysis: n.19G>A, n.104T>C and n.116_127del were all Tier 1 variants, while n.159_176dup and n.182_190del overlapped Tier 1 variants. Furthermore, n.100T>G and n.104T>G were Tier 2 variants. Neither of the other two variants (n.31G>A and n.148C>A) were observed in the NGRL. All variants in the replication collections were inherited in trans except for one variant, which appeared de novo on the paternal allele (**Fig. 3b**); none were observed in homozygosity in gnomAD v4. The HPO terms assigned to the seven cases revealed a broad consistency with the terms of Tier 1 and Tier 2 cases, including ’Global developmental delay’ and ’Seizure’ (**Fig. 3c**). There were no reports of NDD-related traits in pedigree members carrying only one of the compound heterozygous *RNU2-2* variants present in the affected relatives.

**Fig. 3.**
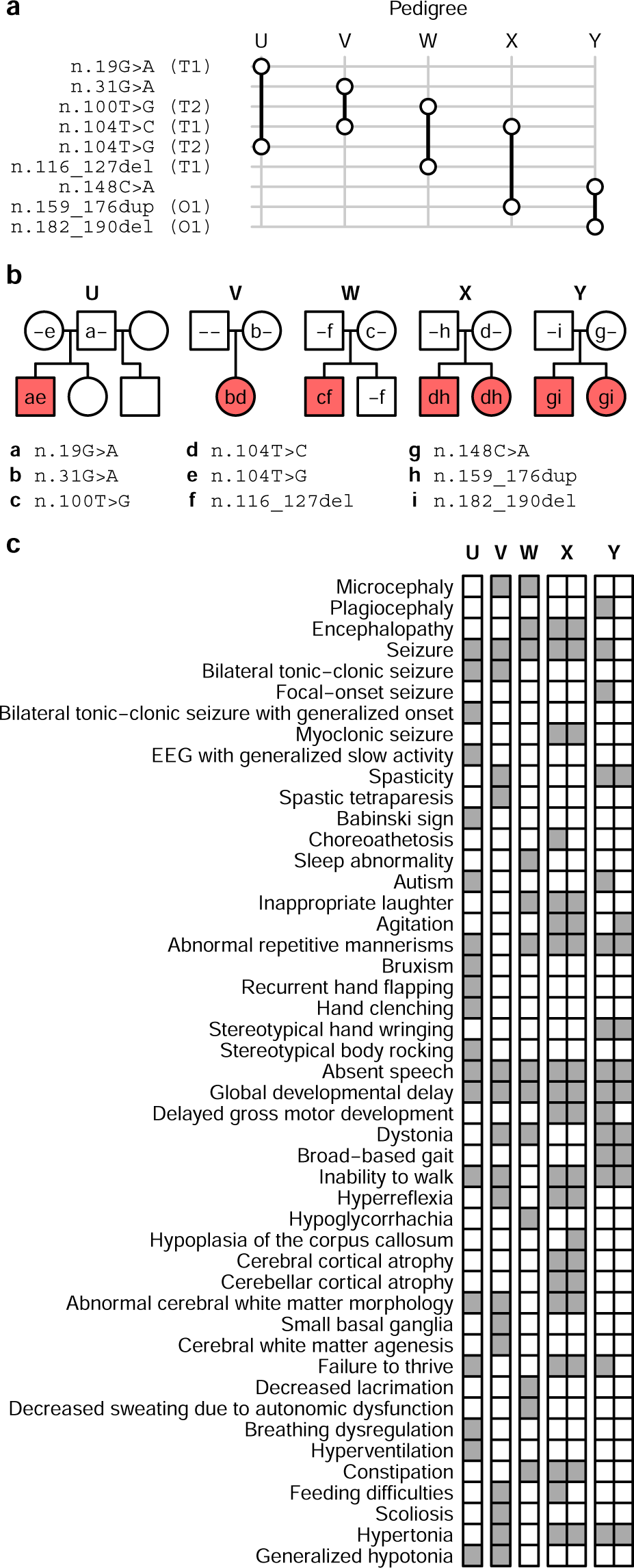
Replication findings. **a**, Variant by pedigree grid for affected pedigrees identified in the UDN (U, V, W and Y) and in the UPP (X) collections. T1: Tier 1 variant; T2: Tier 2 variant; O1: indel overlapping a Tier 1 variant. The symbol representations match those in Fig. 2a. **b**, Pedigrees in the replication collections. The symbol representations match those in Fig. 2c. **c**, Grid of HPO terms assigned to the cases in the replication pedigrees. The HPO terms for the affected brother (left) and sister (right) in pedigree X are shown as two attached columns.

Detailed clinical vignettes for eight cases from six families (pedigrees F, U, V, W, X and Y) are provided in **Supplementary Note** and summarized in **Supplementary Table 1**. These indicate that the recessive NDD caused by biallelic variants in *RNU2-2* was typically first recognized in infancy or early childhood. Pregnancy and perinatal histories were usually unremarkable, although there were occasional reports of jaundice and prematurity. Presenting features included hypotonia, global developmental delay and isolated expressive/receptive language impairment. The phenotype spanned from mild learning disability with autism spectrum disorder to severe, progressive epileptic encephalopathy. Motor impairment ranged from normal ambulation to complete non-ambulation. There were also reports of dystonia, spasticity, and choreoathetosis. Speech was typically reported as being limited to a few words or absent. Epilepsy was common, often beginning in infancy or early childhood, and could evolve to drug-resistant Lennox–Gastaut syndrome; EEGs showed diffuse slowing and epileptiform discharges. Brain MRI scans were often normal early in life but could later show cerebral and cerebellar atrophy or white matter changes (see **Extended Data** Fig. 4). Severely affected cases developed scoliosis, feeding dependence, and autonomic instability. Premature death occurred in one case resulting from prolonged seizures and respiratory compromise. Milder cases remained stable into adolescence or adulthood but with persistent cognitive and behavioural difficulties. In participants with available clinical images, a distinctive or consistent facial gestalt was not evident, although some individuals were dysmorphic. For example, the proband from pedigree V had a short philtrum and prominent central incisors/macrodontia (**Fig. 4**).

**Fig. 4.**
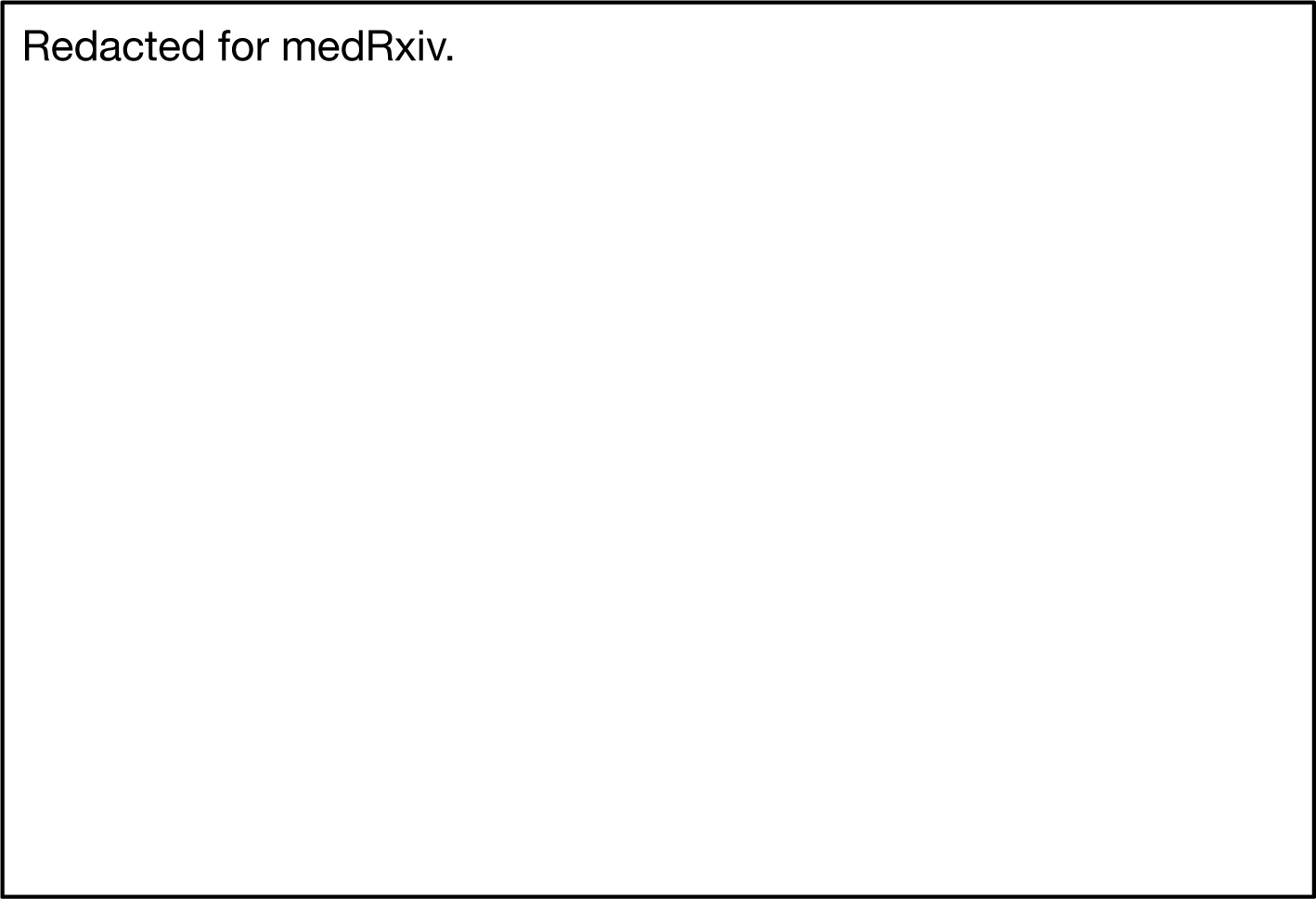
Clinical photographs. Among the seven participants from five families with available photographs, no obvious characteristic facial gestalt was observed; however, some individuals exhibited dysmorphic features.

In the 100KGP, recessive *RNU2-2* syndrome was, by a significant margin, the most prevalent etiological gene for a recessive NDD, accounting for the only biallelic cases amongst the top 30 most prevalent etiological genes for NDDs (**Extended Data** Fig. 5). No other gene apart from *RNU2-2* accounted for more than four biallelic NDD probands in the 100KGP. The cases with recessive *RNU2-2* syndrome amounted to between 36% (Tier 1 cases only) and 62% (Tier 1 and Tier 2 cases) of the number of ReNU cases. Recessive *RNU2-2* syndrome accounted for between 7.6% (Tier 1 cases only) and 13.1% (Tier 1 and Tier 2 cases) of all NDD probands with a biallelic genetic diagnosis in the 100KGP. In the GMS, which has less ascertainment bias due to prior testing, the proportions were 6.7% and 9.8%, respectively. In the 100KGP and the GMS, which contains nine cases with dominant *RNU2-2* syndrome due to mutations at n.4 or n.35, seven Tier 1 pedigrees and all 13 Tier 2 pedigrees with biallelic variants in *RNU2-2* were non-consanguineous. It is therefore very likely that, even in populations without any consanguineous relationships, recessive *RNU2-2* syndrome is more prevalent than the dominant form. Furthermore, the recessive form is liable to affect multiple siblings within pedigrees, further boosting its prevalence.

The U2 class of snRNAs encoded by *RNU2-2* or its canonical counterpart *RNU2-1*, interact through branch point binding sites (BPbs) with intronic branch sites in protein coding pre-mRNAs. U2 snRNAs also interact with multiple other snRNAs and RNA binding proteins within the major spliceosome which, collectively, maintain quaternary structure and function during the transition from the A to the B spliceosome complex, which has catalytic activity for intron excision^18,19^. Dominant *RNU2-2* syndrome results from variants predicted to disrupt intermolecular interactions with U6 or with the branch sites of introns^4^. By contrast, most variants that cause recessive *RNU2-2* syndrome were predicted to disrupt intra-molecular interactions, in most cases by abolishing Watson-Crick pairings within the U2-2 stem loop motifs (**Fig. 5**). The stem loops have ribonucleoprotein binding functions but also undergo dynamic remodelling during spliceosome assembly that is necessary for the correct presentation of the BPbs for intron recognition and for the acquisition of spliceosome catalytic activity. Other variants were located within the BPbs itself (n.31G>A, n.39T>C, n.43T>C and n.45C>T) or were within the site that interacts with the Sm-type heteroheptametric ring (n.100T>G, n.104T>G and n.104T>C). Structural modelling of the Tier 1 variants predicted significant destabilization of stem loop motifs or intermolecular interactions that likely disrupt multiple aspects of spliceosome function (**Supplementary Table 2**).

**Fig. 5.**
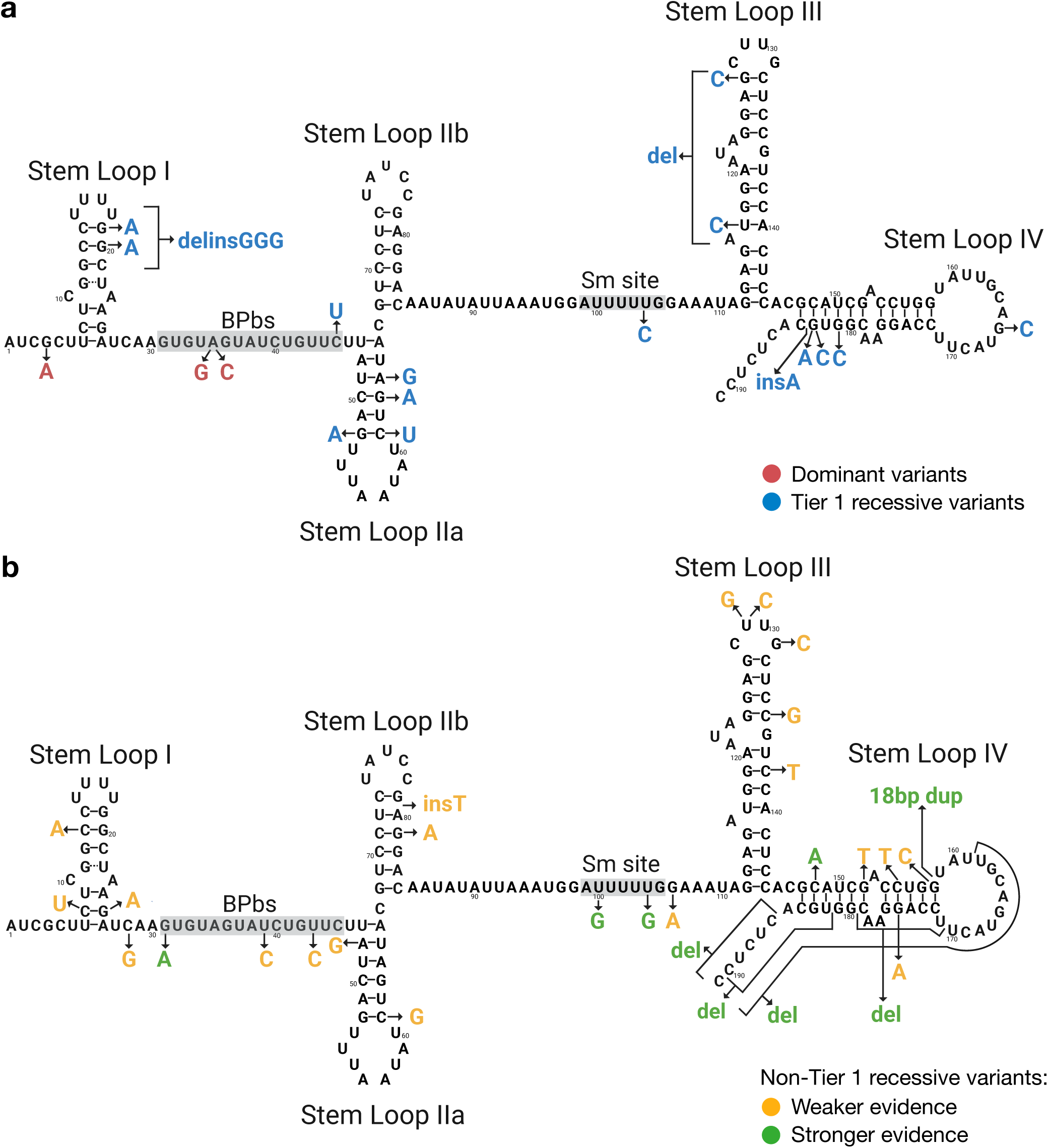
Locations of the possibly pathogenic variants in *RNU2-2*. **a,** Schematic of the secondary structure of human U2-2 snRNA with ribonucleotide numbering according to the canonical *RNU2-2* reference sequence ENST00000410396. Stem loops 1, IIa, IIb, III and IV, the branch-site binding site (BSbs) and the Sm interacting site (Sm site) are taken from the U2-1 structure described in Feltz et al.^18^. The ribonucleotide substitutions at positions 4 and 35 previously reported to be causal of dominant *RNU2-2* syndrome are indicated in red^4,6^. Tier 1 variants, which are likely to be responsible for recessive *RNU2-2* syndrome, are indicated in blue. Note that n.183_184 insA arose on a haplotype containing n.67C>T but, because n.67C>T is much more common and is outside any domains or stem loops, only the change in ribonucleotide sequence induced by n.183_184 insA is shown. **b**, Schematic showing variants that are not Tier 1 variants. Indels that overlap a Tier 1 variant or a variant in the replication collections and Tier 2 variants that are observed in the replication collections are shown as having stronger evidence (green) of pathogenicity. The remainder are shown having weaker evidence (amber) of pathogenicity.

Given the well known function of U2 snRNAs within the major spliceosome, we speculated that cells from cases with *RNU2-2* syndrome might exhibit aberrant splicing. To explore this hypothesis, we analyzed whole blood RNA-seq alignments from three unrelated cases and 497 unrelated NGRL participants with unexplained NDDs (we previously showed that *RNU2-2* is robustly expressed in blood^4^). This analysis did not reveal differential gene expression of note, differential splice junction usage or any pattern of aberrant splicing in the cases (**Extended Data** Fig. 6). Although this may be partly attributable to a lack of power, our prior observation of similar results obtained by analyzing five cases with dominant *RNU2-2* syndrome^4^ suggests that transcriptomic analysis of other tissue types will be required to understand the molecular pathophysiology of both *RNU2-2* disorders.

Having previously identified statistical associations between dominant NDDs and variants in *RNU4-2* and *RNU2-2*^3,4^, we have now demonstrated a statistical association with a third recessive NDD caused by different variants in *RNU2-2*. Aggregating over the monoallelic and both tiers of biallelic cases in the 100KGP, the number of cases with an *RNU2-2* syndrome, which a family foundation^20^ has recently named ReNU2, is 81% that of cases with ReNU. Overall, we identified 23 Tier 1 cases from 18 pedigrees (**Fig. 2c**), 15 Tier 2 cases from 13 pedigrees (**Extended Data** Fig. 1) and 7 replication cases from 5 pedigrees (**Fig. 3b**), giving a total of up to 45 cases with a credible diagnosis of recessive ReNU2. Interestingly, recent unpublished work from a different group points to the existence of a recessive NDD due to variants in *RNU4-2* as well^21^. However, that disorder is likely to be much rarer than recessive ReNU2 given that in the NGRL there are only seven previously unexplained NDD cases with biallelic rare variants in *RNU4-2* but 50 cases with such variants in *RNU2-2* (of which 38 are Tier 1 or Tier 2 cases).

The dominant ReNU and ReNU2 disorders have very limited genetic heterogeneity, as they are caused by variants in very narrow regions within their respective genes. In contrast, the recessive ReNU2 disorder is caused by a wide array of variants affecting various stem loops and both protein and intron branch site binding domains of U2-2 snRNA. This may explain the greater phenotypic heterogeneity among cases with recessive ReNU2 syndrome compared to those with dominant ReNU or ReNU2 syndromes. For example, of the cases for which detailed phenotype data were available, the case in pedigree F had a particularly mild phenotype, with no seizures, only slight dysmorphic features and mild intellectual disability. One of her two variants (n.20G>A) was present in other cases with a severe phenotype, so is probably highly impactful. Her other variant (n.61C>T) disrupts a duplex (r.52–r.61) at the base of stem loop IIa, which is also perturbed in a severely affected case (in pedigree B) by n.52G>A, the equivalent of which has been experimentally verified to be destabilizing in yeast^22^ (**Fig. 5**). However, the classic wobble base pairing G–U at r.52–r.61 in pedigree F might be more stable than the A–C pairing in pedigree B, which would explain the milder phenotype. The example underscores the need to gather larger case collections to refine the relationships between variants in *RNU2-2*, many of which have never been seen in homozygosity, and the spectrum of phenotypes associated with recessive ReNU2 syndrome.

## Data Availability

Genetic and phenotypic data for the 100KGP study participants, the 100KGP Pilot study participants and the GMS participants are available through the Genomics England Research Environment via the application at https://www.genomicsengland.co.uk/join-a-gecip-domain. WGS data in the NGRL were obtained for 78,132 100KGP participants, 4,054 100KGP Pilot participants and 32,030 GMS participants (v3) HPO phenotype data in the NGRL were obtained from the 'rare_diseases_participant_phenotype' table (Main Programme v14), 'observation' table (GMS v3) and 'hpo' table (Rare Diseases Pilot v3); Specific Disease class data from the 'rare_diseases_participant_disease' table (Main Programme v13); ICD10 codes from the 'hes_apc' table (Main Programme v13); pedigree information from the 'rare_diseases_pedigree_member' table (Main Programme v13), 'referral_participant' table (GMS v3), and 'pedigree' table (Rare Diseases Pilot v3); explained/unexplained status of cases from the 'gmc_exit_questionnaire' table (Main Programme v18, GMS v3); consanguinity status of families from the 'participant' table (Main Programme v14). Ensembl v104 (http://may2021.archive.ensembl.org/index.html), gnomAD v3.0 (https://gnomad.broadinstitute.org/) and CADD v1.6 (https://cadd.gs.washington.edu/), were used for transcript selection and variant annotation. The most recent version of gnomAD (v4.1.0) was also used where specified. UDN genomes are available to approved researchers via dbGaP(ref. phs001232.v7.p3). The main website of the UDN at https://undiagnosed.hms.harvard.edu/about-us/ contains further details and instructions on data access. Data presented in this paper were requested from the Genomics England Airlock on 7 August 2025 at 20:25 BST. The manuscript was submitted to the Genomics England Publication Committee on 15 August 2025 at 23:51 BST and approved for submission on 21 August 2025 at 17:02 BST.

http://may2021.archive.ensembl.org/index.html

https://gnomad.broadinstitute.org/

https://cadd.gs.washington.edu/

https://undiagnosed.hms.harvard.edu/about-us/

## Acknowledgements

This research was made possible through access to data in the NGRL, which is managed by Genomics England Limited (a wholly owned company of the Department of Health and Social Care). The NGRL holds data provided by patients and collected by the NHS as part of their care and data collected as part of their participation in research. The NGRL is funded by the National Institute for Health Research and NHS England. The Wellcome Trust, Cancer Research UK and the Medical Research Council have also funded research infrastructure. This work was supported by the Italian Ministry of Health (RCR-2022-23682289, PNRR-MR1-2022-12376811 and RF-2021-12374963 to M.T.). Research reported in this publication was supported by the National Institute of Neurological Disorders and Stroke of the National Institutes of Health under Award Number U01NS134358. D.G., A.M., and E.T. were supported by the National Heart, Lung and Blood Institute of the National Institutes of Health under Award Number R01HL161365. The content is solely the responsibility of the authors and does not necessarily represent the official views of the National Institutes of Health.

## Author contributions

D.G. conducted statistical and bioinformatic analyses and cowrote the paper. R.M. was the lead UDN clinical analyst and cowrote the paper. J.L. modeled the molecular impacts of variants. M.B. prepared the phenotype table. A.B., L.C. and C.M. were responsible for molecular genetic analyses in Italy. E.S.B., D.E.B. and M.W. consented and provided detailed clinical data. M.T. coordinated the Italian UPP study. J.A.B. and M.T.W. supervised the Stanford UDN study. A.D.M. coordinated clinical contacts, provided clinical and biological interpretation, and cowrote the paper. E.T. oversaw the study and cowrote the paper.

## Methods

### Enrollment

The enrollment criteria for participants in the NGRL are available from the Genomics England website^23^. The UDN enrollment criteria, detailed in the UDN Manual of Operations^24^, specify eligibility for individuals with unexplained conditions despite prior evaluation and sufficient clinical data. UDN cases are triaged, and clinical sites perform multidisciplinary evaluations, including genomic testing, to advance diagnosis and discovery.

### Genetic association analysis

We built a Rareservoir^15^ comprising 110,009 participants in the NGRL enrolled into the 100,000 Genomes Project (100KGP), which included 78,195 study participants, and the Genomic Medicine Service (GMS), which included 31,814 study participants. The Rareservoir only contained genotypes for rare variants, defined as those being absent from gnomAD or having a gnomAD PMAF score^15^ ≥ 1 (i.e., likely to have a minor allele frequency < 1/1,000 in all gnomAD populations). As the 100KGP and the GMS used different bioinformatic pipelines, we only analyzed variants in regions in which at least 95% of samples from each collection passed quality control according to the FILTER column in the gVCF files.

Relatedness estimates were available on the NGRL within the 100KGP but not between the 100KGP and the GMS nor within the GMS. To identify closely related pairs of participants where at least one participant was in the GMS, we applied the following computationally efficient approach. For each such pair of participants (𝑎, 𝑏), we declared the pair closely related if 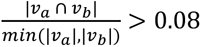, where 𝑣 denotes the set of participant 𝑥’s autosomal rare variants that are present in gnomAD and have a PMAF score ≥ 2 (i.e., they are likely to have a MAF < 1/10,000 in all gnomAD populations). We merged pairs of families determined by the ’rare_disease_pedigree_member’, ’referral’ and ’referral_participant’ tables if they contained closely related individuals recursively until all closely related individuals were assigned to the same families. This approach tends to overmerge (i.e., it may occasionally assign individuals who are not closely related to the same family), but it is conservative in that it might reduce power but not generate false positive associations. To identify a maximal set of unrelated participants for use in our statistical association analysis we started with the complete set of participants and repeatedly removed the person to whom the greatest number of remaining people were closely related according to the definition above.

We performed an initial genetic association analysis of non-coding genes (including pseudogenes) using BeviMed^16^ as described previously^4^. The NDD case group comprised 12,776 unrelated participants (i.e., belonging to different families) who had the NDA HPO term (after imputing the “Intellectual disability” HPO term into cases in the 100KGP Specific Disease group “Intellectual disability”) or had the GMS referral code for “Intellectual disability.” The controls comprised 56,064 unrelated participants who did not have an NDD according to those criteria. Of the 12,776 participants in the case group, 2,886 had been previously solved through pathogenic or likely pathogenic variants. Cases explained by variants in a given gene were reassigned to the control group in the genetic association analysis for that gene. The initial BeviMed runs yielding a PPA>0.5 were re-run with the following adaptations. Firstly, the variants within the associated genes were phased using whatshap v2.7^17^. Secondly, the BeviMed model was adapted so that a configuration of alleles under the recessive model was considered pathogenic only if both haplotypes contained a pathogenic allele (i.e., having two pathogenic alleles in cis would not be considered a pathogenic configuration of alleles). BeviMed 7.0 implements this new functionality.

### Determining consanguinuity

The ‘participant’ table in the NGRL was used to determine consanguinity. Note that pedigrees J and R were labelled as consanguineous even though their consanguinity status was absent because their respective cases had homozygous to heterozygous genotype count ratios for variants with a PMAF score ≥ 2 in the top 1% of all participants, which was strongly indicative of consanguinity.

### Phenotypic homogeneity analysis

To assess the phenotypic homogeneity of the 12 Tier 1 cases in the 100KGP, we computed a phenotype homogeneity score for that group with respect to unexplained NDD probands in the 100KGP. We calculated this score using the ‘get_sim_grid’ and ‘get_sim_p’ functions from the ontologySimilarity R package,^25^ as previously described^3^. We then obtained a Monte Carlo *P*-value as the proportion of random sets of 12 NDD probands having a homogeneity that was greater than or equal to the homogeneity score of the group of 12 Tier 1 probands.

### Analysis of HPO terms

To identify enriched or depleted HPO terms among the 12 NDA-annotated Tier 1 probands in the 100KGP compared with other unexplained NDD probands, we computed *P*-values of association using Fisher’s two-sided exact test. We only tested enrichment for terms attached to at least three of the 12 Tier 1 probands and which belonged to the set of non-redundant terms at each level of frequency among the probands. To account for multiple comparisons, we adjusted the *P*-values by multiplying them by the number of tests. An adjusted *P*-value < 0.05 was deemed statistically significant. To visualize both common and distinctive HPO terms for biallelic *RNU2-2* cases, we selected terms that were either statistically significant or present in at least 50% of the cases, removed redundant terms at each level of frequency among the 12 probands, and arranged the terms along with a non-redundant set of ancestral terms as a directed acyclic graph of is-a relations. These analyses were conducted using the ontologyX R packages^25^.

### Modeling free energies of association

Unless otherwise stated, we modelled the effects of mutations energetically with ViennaRNA v2.7.0^26^, using the secondary structure shown in **Fig. 5** and the U2-2 sequence as reference wild type. We used the ViennaRNA RNA.fold_compound.eval_structure function to obtain the ΔG of folding for both wild type and mutant U2-2 snRNA, allowing us to calculate the change in stability, ΔΔG, upon mutation, in kcal/mol (positive values indicating relative destabilization). We analyzed spliceosome structures using the RSCB^27^ online PDB viewer. We used Chimera-X^28^ to assess the effects of mutations on U2-2 snRNA-protein interactions (e.g., to evaluate potential changes in hydrogen bonds).

### Gene expression and splicing analysis

We performed QC on RNA-seq data derived from the whole blood of 5,546 participants in the NGRL as follows. Based on visual inspection of QC parameter distributions, we filtered out samples with a percentage of RNA fragments larger than 200 bases (as measured by an Agilent Tapestation 4200) ≤ 65%, a total read count outside the range (108M, 592M), a genome mapping rate < 0.85 or a high-quality read rate < 0.9 (where reads were deemed to be of high quality if they aligned as proper pairs, had fewer than seven mismatches and had a mapping quality ≥ 60). After QC filtering, 5,165 samples remained for analysis, including three Tier 1 cases. We selected 500 samples for differential gene expression and splice junction usage analysis by taking the three Tier 1 cases and 497 samples selected at random from those passing the QC criteria, and which belonged to NDD probands who are presently unexplained. We used DESeq2^29^ (version 1.44) to conduct differential gene expression analysis using the transcript quantifications generated by the Salmon software^30^ and aggregated them by gene using the ’tximport’ BioConductor package^31^. To perform differential splicing analysis, we computed a normalized junction usage for each junction and sample by dividing the number of uniquely aligned reads supporting that junction by the total number of uniquely aligned reads supporting any junction within the sample. To assess whether at least one junction was significantly overused by the three Tier 1 cases, we calculated the sum of ranks for the three cases among the 500 of the normalized usage (ranked from high to low) of each junction and took the lowest value across all junctions (we analyzed 1,311,470 junctions that were supported by at least one spliced read in at least three of the 500 samples). We then computed a one-sided permutation *P*-value by computing the sum of ranks for 500 random sets of three samples. Analogously, to determine whether any one junction was significantly underused by the three Tier 1 cases, the same procedure was used, except that (i) the junctions were ranked by normalized usage from low to high instead of high to low, and (ii) we only included splice junctions observed in at least 95% of the 500 samples, comprising 110,892 junctions. To assess whether a group of splice junctions was significantly overused or underused by the Tier 1 cases, we computed the mean normalized usage over cases and junctions in the group and compared that value to the distribution of such values obtained by random permutation of case labels. The proportion of random sets yielding a greater value than that obtained for the Tier 1 cases gave the over-usage *P*-value while the proportion of random sets yielding a lower value than that obtained for the Tier 1 cases gave the under-usage *P*-value. We applied this analysis to junctions grouped by: dinucleotide pairs at the splice sites, quantile of GC content in the region encompassed by the splice junction, and quantile of splice junction length.

### Ethics

Participants of the 100KGP and the GMS were enrolled to the NGRL under a protocol approved by the East of England–Cambridge Central Research Ethics Committee (ref: 20/EE/0035). We obtained written informed consent to publish additional clinical data from a subset of the affected cases in the NGRL following local best practices. The UPP study was approved by the Institutional Ethical Committee of the Ospedale Pediatrico Bambino Gesù in Rome (refs. 2072_OPBG_2020, PNRR-MR1-2022-12376811, 1702_OPBG_2018 and RF-2021-12374963). Ethical and research approvals for participants in the UDN were obtained from the Stanford University Institutional Review Board (protocol 47026) and the National Human Genome Research Institute Institutional Review Board (protocol 15-HG-0130). All participants provided written informed consent.

## Code availability

Software packages rsvr 1.0, bcftools 1.16, samtools 1.9 and perl 5 were used to build the 100KGP Rareservoir. The Rareservoir software is available from https://github.com/turrogroup/rsvr. All R packages listed in the manuscript are available via the Comprehensive R Archive Network site (https://cran.r-project.org/) or Bioconductor (https://bioconductor.org). The ViennaRNA 2.6.4 and whatshap 2.6 packages are available from the Python Package Index (https://pypi.org).

---

**Extended Data Fig. 1.**
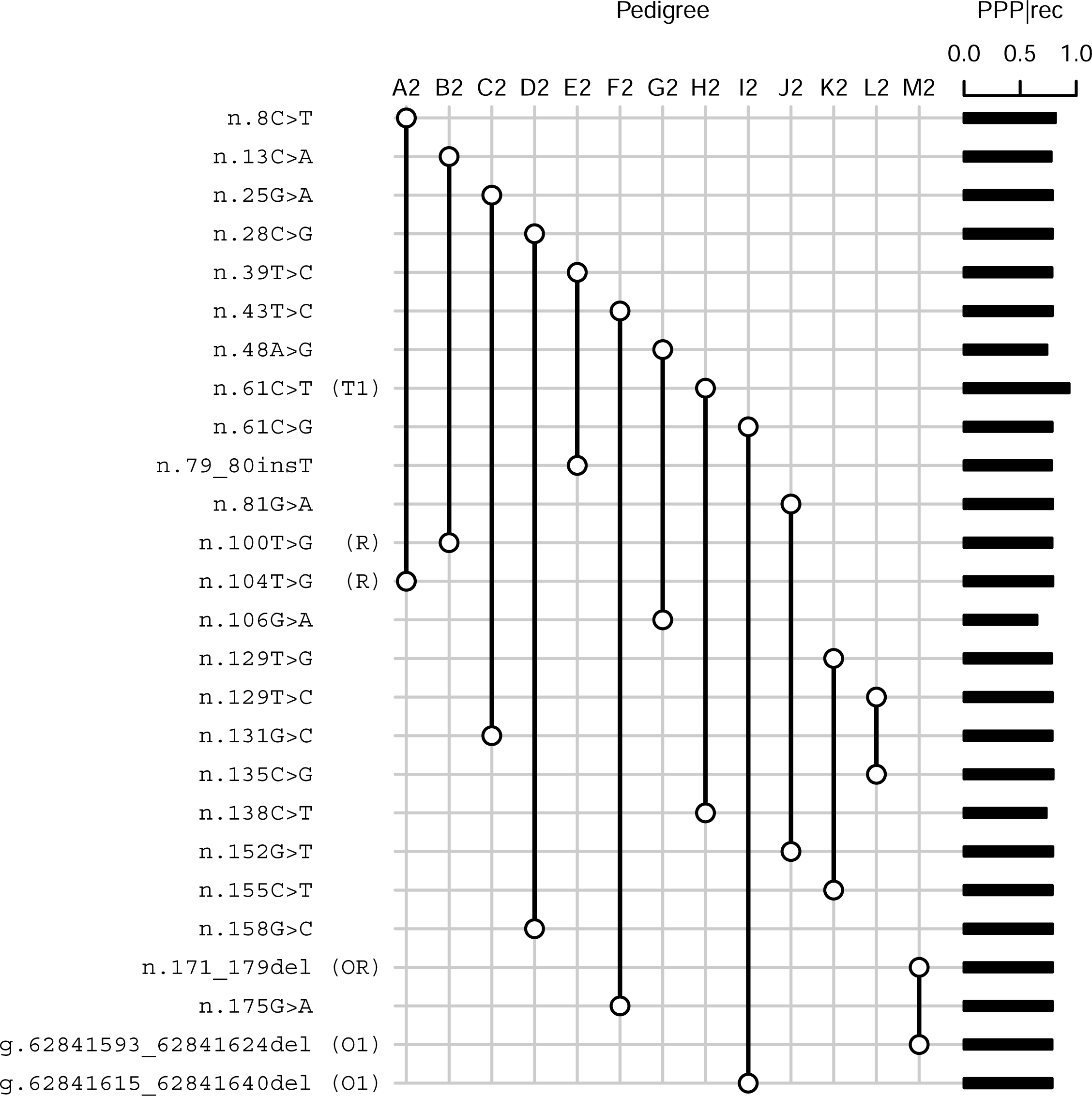
Variants in Tier 2 cases. Variant by pedigree grid illustrating the variants and genotypes corresponding to the Tier 2 cases. A pair of empty circles connected by a line denotes heterozygous genotypes in trans. The PPP|rec values for each variant are shown as bars on the right. The compound heterozygous variants in the probands of D2 and M2 were present in one affected sibling of each family and there were no examples of non-cosegregation. T1: Tier 1 variant; O1: indel overlapping a Tier 1 variant; R: variant observed in the replication collection; OR: indel overlapping a variant in the replication collection.

**Extended Data Fig. 2.**
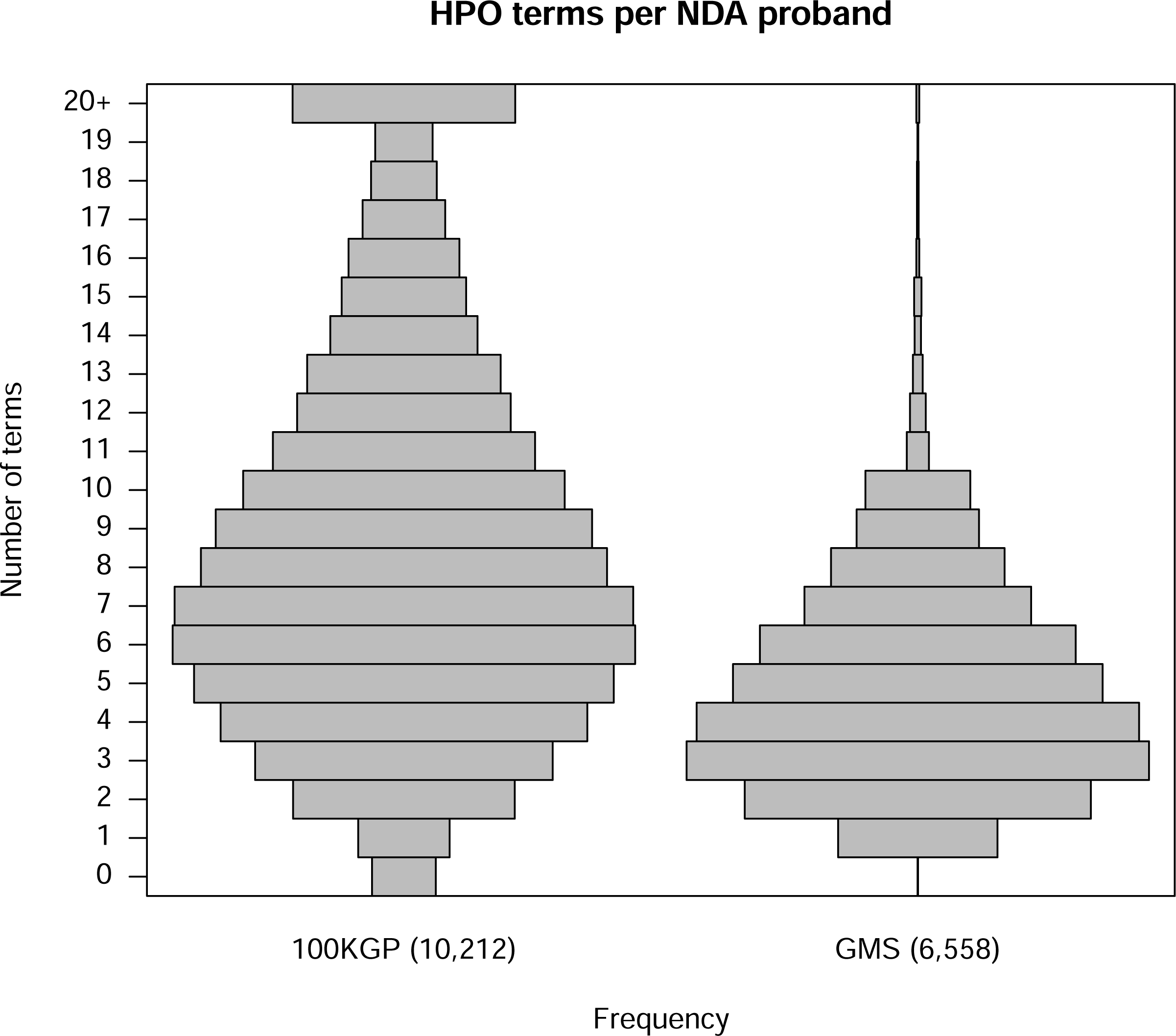
HPO coding in the 100KGP and the GMS. Histograms showing the number of HPO terms assigned to probands with the ’Neurodevelopmental abnormality’ (NDA) HPO term in the 100KGP and the GMS.

**Extended Data Fig. 3.**
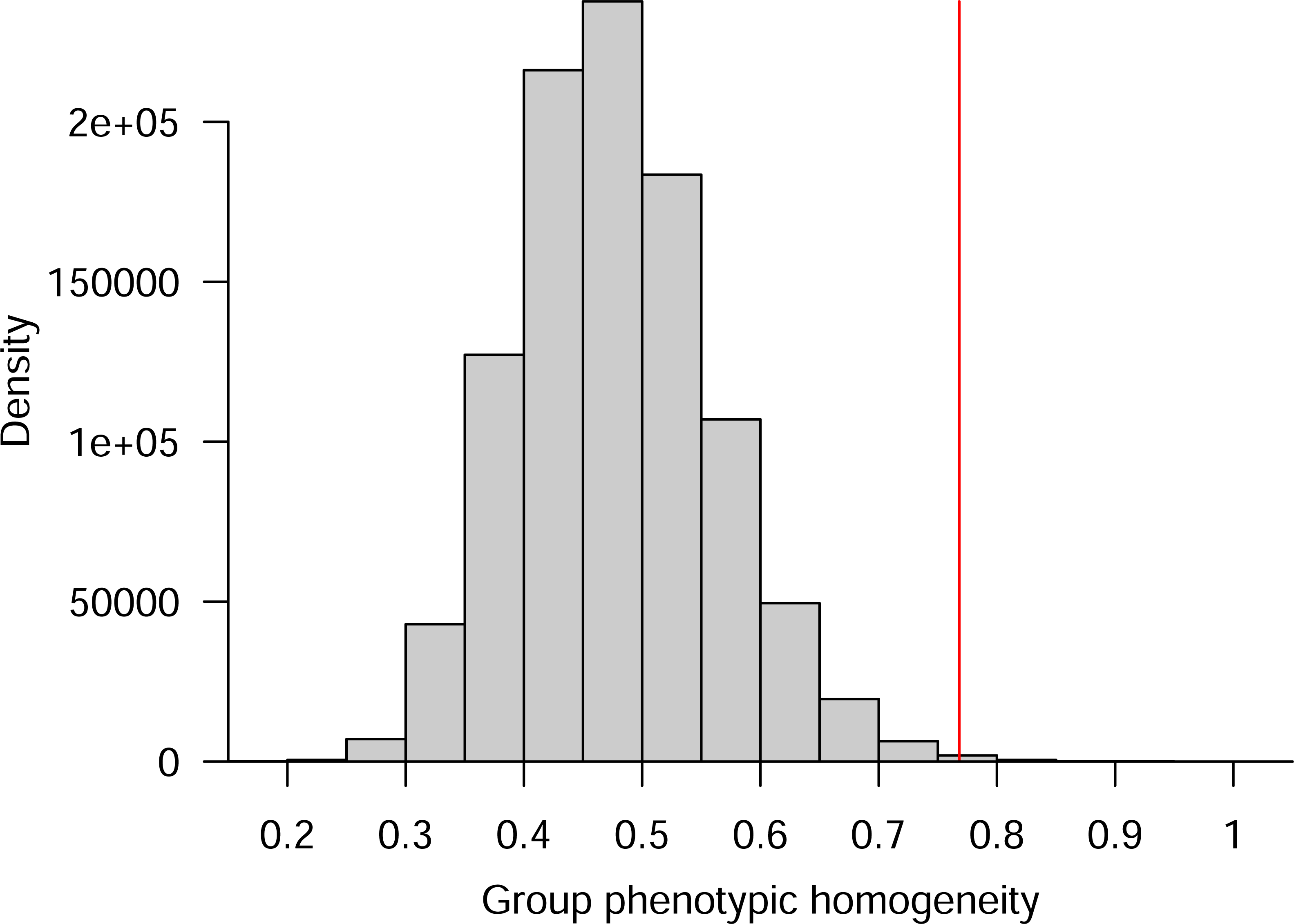
Phenotypic similarity of Tier 1 cases. Distribution of phenotypic homogeneity scores for 100,000 randomly selected sets of 12 probands chosen from 9,112 NDD probands. The score corresponding to the 12 Tier 1 cases with recessive *RNU2-2* syndrome is indicated with a red line.

**Extended Data Fig. 4.**
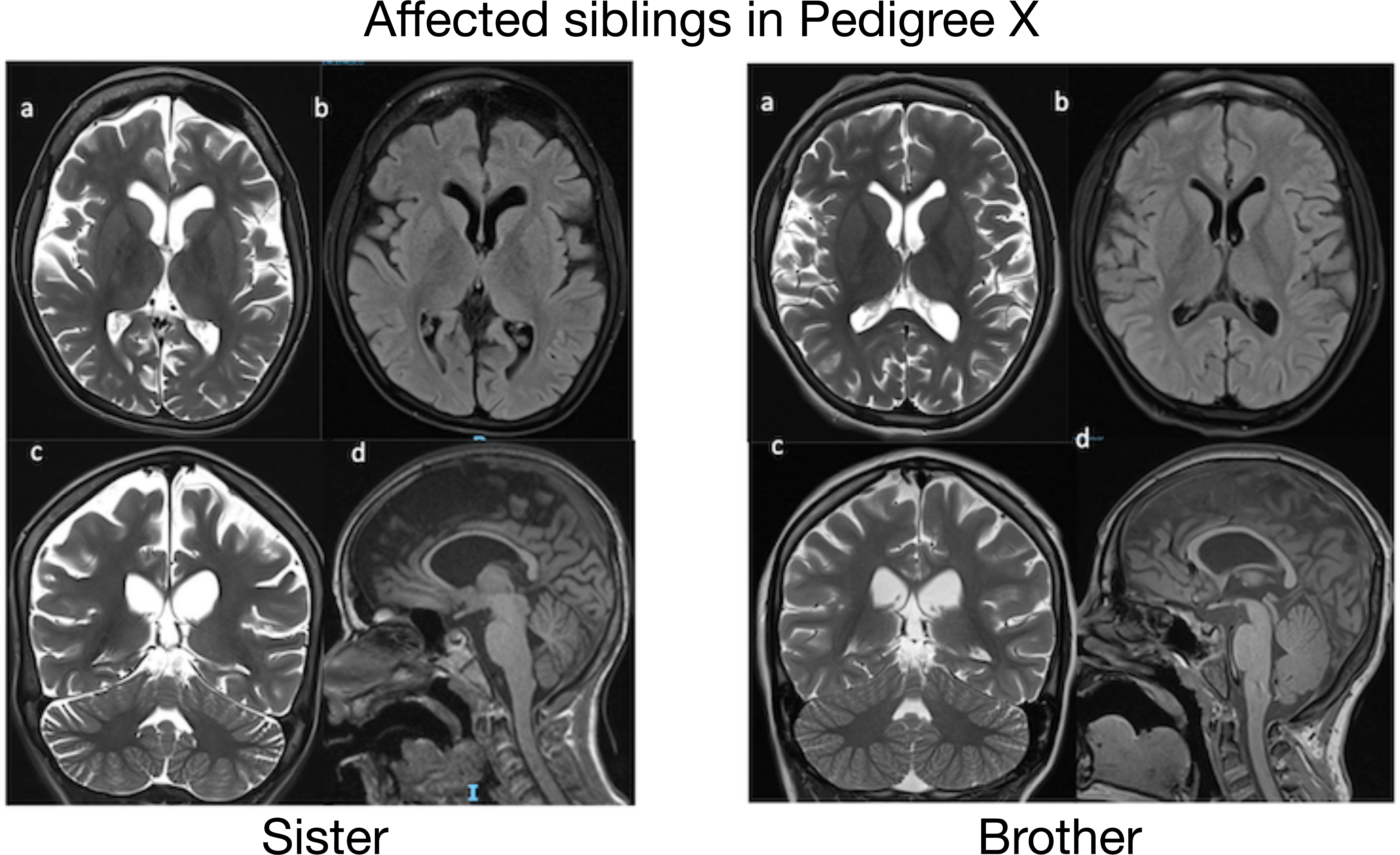
Brain MRI for two affected siblings in pedigree. **X.** The upper row shows T2-weighted (**a**) and FLAIR-weighted (**b**) axial images with a section through the basal ganglia. There are increased subarachnoid spaces suggesting cortical atrophy together with moderate ventricular enlargement. There is also a slight hyperintensity of the white matter. The lower row shows a T2-weighted coronal image (**c**) featuring signs of brain cortical atrophy and diffusely enlarged cerebellar interfoliar spaces, and a T1-weighted sagittal image featuring the corpus callosa (**d**).

**Extended Data Fig. 5.**
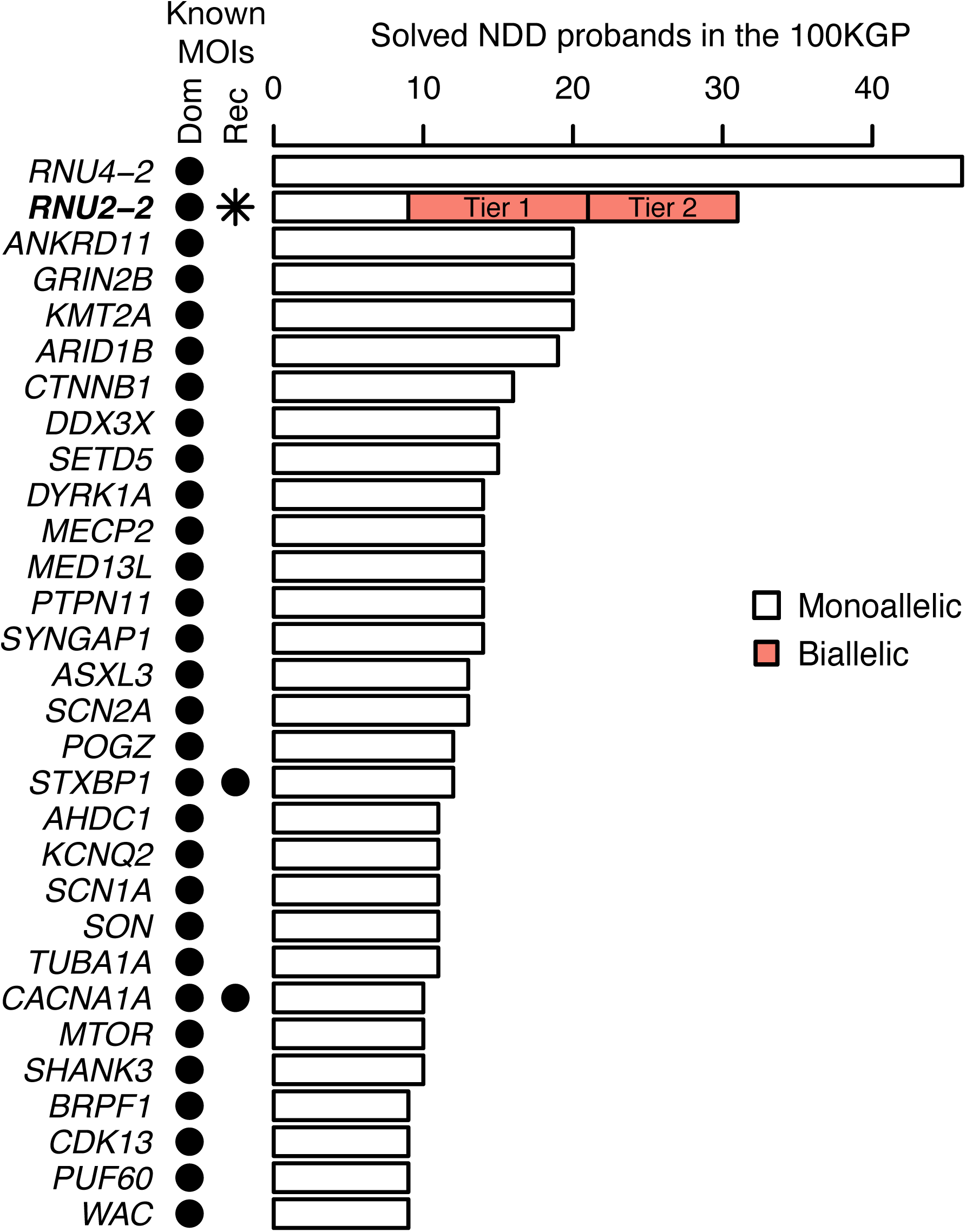
Prevalence in the 100KGP. Of the 9,112 unrelated NDA-coded cases in the 100KGP, the numbers solved through pathogenic or likely pathogenic variants in a gene are shown, provided at least nine cases were diagnosed. The bars have been partitioned in order to show monoallelic and biallelic cases separately. For dominant *RNU4-2* and *RNU2-2* disorders, the numbers of NDA-coded cases with causal variants reported in Greene et al. 2024 and Greene et al. 2025 are shown, respectively, because the corresponding clinical reporting has still not percolated to the ’gmc_exit_questionnaire’ table in the NGRL. The number of cases with recessive *RNU2-2* syndrome is partitioned into high-confidence Tier 1 cases and lower-confidence Tier 2 cases in the 100KGP. Known modes of inheritance based on PanelApp data are shown as columns of points. The proposed recessive mode of inheritance for *RNU2-2* is indicated with an asterisk.

**Extended Data Fig. 6.**
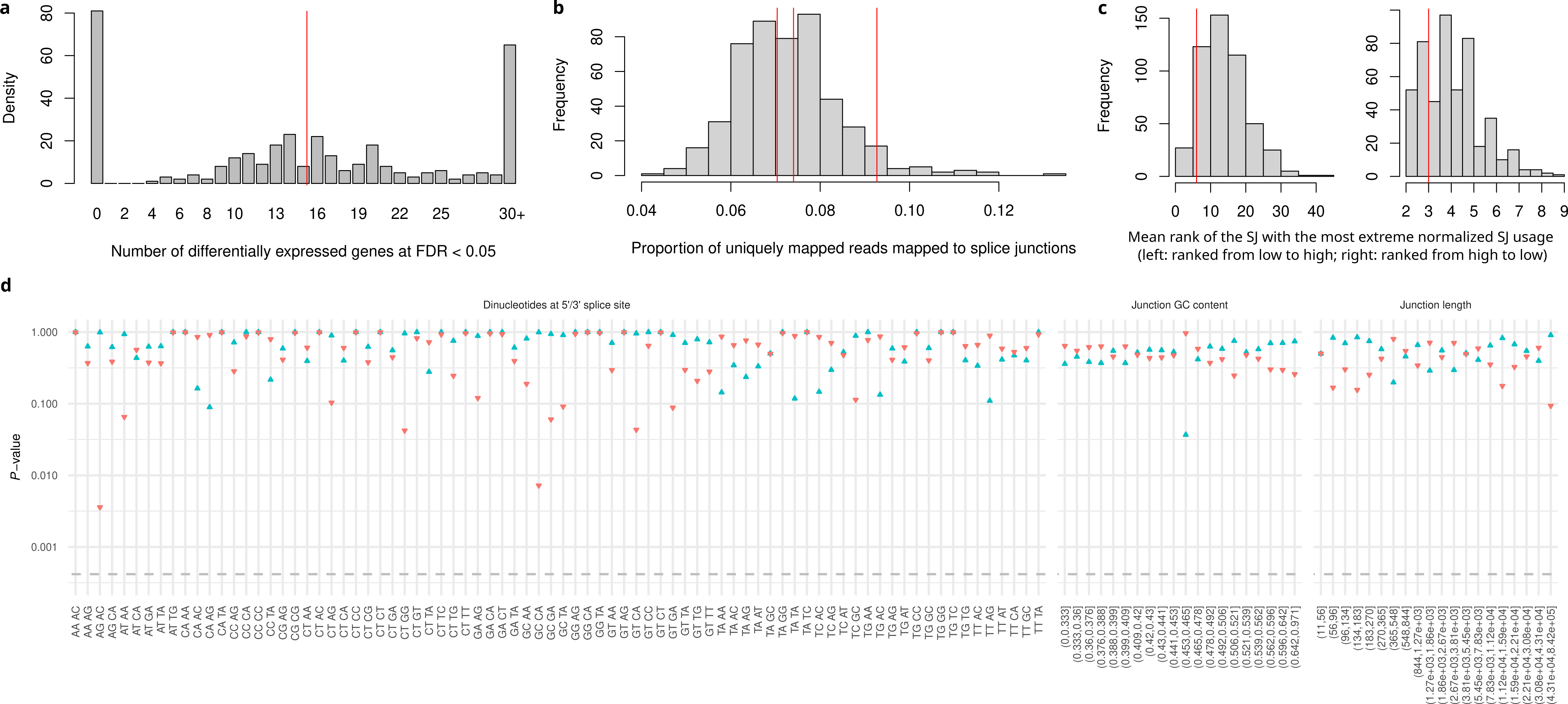
RNA-seq analysis of whole blood. **a,** Histogram of the number of differentially expressed genes, controlling FDR at 0.05 with the Benjamini-Hochberg procedure, for randomly selected sets of three from 500 RNA-seq samples (comprising the biallelic cases in pedigrees F, Q and P, and 497 unexplained unrelated NDD cases without variants in *RNU2-2*). The number of such genes for the three recessive *RNU2-2* cases is shown with a red line. **b**, Histogram of the proportion of uniquely aligned RNA-seq reads that contain a splice junction in the 500 RNA-seq samples. The proportions corresponding to the samples from the three biallelic cases are shown with red bars. **c**, Histogram of the mean (over randomly selected sets of three samples) rank of normalized splice junction (SJ) usage of the splice junction with the lowest mean rank after ranking from low to high usage (left) and from high to low usage (right) and assigning the maximum rank in the event of ties. The two red lines correspond to the lowest mean ranks obtained for the three biallelic *RNU2-2* cases. **d**, One-sided *P* values obtained by permutation of case labels within the 500 samples for the lowness of the sum of ranks of normalized numbers of reads supporting groups of splice junctions ranked from high to low (the upward facing blue triangles) and low to high (the downward facing red triangles), assigning the maximum rank in the event of ties. The splice junctions were grouped by: dinucleotide pairs at the splice sites (for *N*≥5), quantile of GC content in the region encompassed by the splice junction, and quantile of splice junction length. The dashed line at y = 0.05/120 indicates the *P* value significance threshold to control the family-wise error rate at 0.05.

The content below has been redacted for medRxiv.

**Supplementary Note Clinical vignettes for cases with recessive *RNU2-2* syndrome.**

**Supplementary Table 1 Table summarizing the phenotypes described in the clinical vignettes.**

**Supplementary Table 2 Complete listing of the reported variants in *RNU2-2* and predicted effects of the Tier 1 variants on spliceosome structure and function.**

